# Investigating social deprivation and comorbid mental health diagnosis as predictors of treatment access among patients with an opioid use disorder using substance use services: a prospective cohort study

**DOI:** 10.1101/2022.12.15.22283515

**Authors:** Emma A Adams, Justin C Yang, Amy O’Donnell, Sarah Minot, David Osborn, James B Kirkbride

**Affiliations:** Population Health Sciences Institute, Faculty of Medical Science Newcastle University, Newcastle upon Tyne, UK; Division of Psychiatry, University College London, London UK; Camden & Islington NHS Foundation Trust, London, UK

**Keywords:** cohort, opioid-related disorders, social deprivation, electronic health records, mental health services

## Abstract

**Purpose:** We investigated the role of social deprivation and comorbid mental health diagnoses in predicting re-engagement with substance use services or contact with crisis and inpatient services for individuals with opioid use disorder in secondary mental health care in inner-city London.

**Methods:** We conducted a prospective cohort study which followed individuals diagnosed with a first episode of opioid use disorder who accessed substance use services between September 2015 and May 2020 for up to 12 months, using anonymised electronic health records. We employed Poisson regression and Cox proportional survival analyses to assess associations between exposures and outcomes.

**Results:** Comorbid mental health diagnoses were associated with higher contact rates with crisis/inpatient services among people with opioid use disorder: incidence rate ratios (IRR) and 95% confidence intervals (CI) were 3.37 (2.11-5.53) for non-opioid substance use comorbidity, 7.02 (3.63-13.31) for a single comorbid mental health diagnosis, and 11.68 (7.53-18.71) for multiple comorbid mental health diagnoses. Social deprivation was not associated with contact rates with crisis/inpatient services. Similar patterns were found with time to first crisis/inpatient contact. Social deprivation and comorbid mental health diagnoses were not associated with re-engagement with substance use services.

**Conclusion:** Comorbid substance and mental health difficulties amongst people who use opioids led to earlier and more frequent contact with crisis/inpatient services during the first 12 months of follow up. Given the common co-occurrence of mental health and substance use disorders among those who use opioids, a better understanding of their needs will ensure they are supported in their treatment journeys.

## INTRODUCTION

In 2020, there were an estimated 61.3 million people globally who used opioids in the past year (1). Opioid use and related deaths are leading to some countries experiencing opioid epidemics (2, 3); an estimated 1.2 million people are expected to die from opioids by 2029 in Canada and the USA (2). Although not at the same level, opioid use is still a public health concern in England. Between 2016 to 2017, there were an estimated 261,294 individuals aged 15 to 64 who used opioids in England, 7.4 per thousand of the population (4). Rates of opioid use vary across England, with more deprived regions experiencing greater prevalence compared with less deprived areas (4). Social deprivation is also strongly associated with risk of several psychiatric disorders, including psychotic disorders (5, 6), raising the possibility of syndemic effects, where the co-occurrence of substance use disorders and mental health disorders in the same population (i.e. a syndemic) or same individuals (co-morbidity or dual diagnoses) share a common aetiology. This possibility is reflected in estimates of comorbid opioid use and mental health diagnoses. For example, in England, approximately 57% of people accessing opioid misuse treatment report a co-morbid mental health treatment need (4). This is consistent with estimates from North American studies, which found rates of co-morbid mental health ranged from 45 to 80% among patients seeking buprenorphine or methadone treatment for opioid dependence (7–11).

Opioid treatment can take place within the community, primary care, secondary care, or residential settings (i.e. supported accommodation), in the form of structured prescribing (e.g. buprenorphine or methadone) and/or psychosocial interventions (4). Nonetheless, treatment effectiveness in the community is lower than efficacy report in randomised controlled trials [RCTs]. For example, abstinence rates after three and six months of treatment in England are nearly 20% lower (12) than average abstinence rates found in RCTs for opioid substitution treatment (13). Evidence suggests that several factors contribute to poor treatment outcomes in real-world settings including relapse, drop out from treatment, and death (12). Existing mental health disorders may also affect outcomes following treatment, although evidence is mixed (9, 10, 14). In addition, exposure to deprivation may also affect ongoing engagement with treatment. The limited previous research investigating social deprivation as a predictor of opioid misuse have tended to focus on prescribing, use, access to support, and patient profiles for opioid use (8, 12, 15–18). Other important outcomes, – such as subsequent substance use service engagement, which may indicate ongoing treatment, or rates of subsequent crisis or inpatient care use, which may indicate relapse – have not been investigated to date.

In comparison to RCTs, routine electronic health record (EHR) data present an opportunity to investigate differences in real-world treatment outcomes within entire healthcare systems rather than in controlled, experimental conditions. Here, we used EHR data from a secondary mental health care service provider in inner-London to investigate the association between social deprivation and comorbid mental health diagnoses and subsequent contact with substance use, crisis or inpatient services in patients diagnosed with opioid-related disorders between September 2015 and May 2020. As a secondary outcome, we investigated the association between social deprivation and comorbid mental health diagnoses and time to accessing crisis and inpatient services. We hypothesised that social deprivation would be negatively associated with accessing services, while comorbid mental health diagnoses would be positively associated with accessing services. Further, we hypothesized that multiple comorbid mental health diagnoses would show greater magnitudes of associations.

## METHODS

### Data source

We constructed a cohort of participants treated for a first episode of opioid-related disorder in the Camden & Islington National Health Service (NHS) Foundation Trust (C&I). C&I is part of the publicly funded healthcare system, which provides free secondary mental health services (including substance use services) for a catchment area of approximately 470,000 residents within the two inner-city London boroughs of Camden and Islington. EHR data were made available via a database known as the Clinical Record Interactive Search (CRIS) system (19). CRIS contains anonymised full clinical records and notes – in both structured fields and interrogable unstructured clinical free-text – of all contacts with C&I for over 160,000 mental health service users from 2009 onwards.

### Sample

We included C&I patients residing in the London Boroughs of Camden and Islington with a first episode of care with a substance use service between September 1, 2015, and May 31, 2020 with a recorded F11 (opioid-related disorders) diagnosis. We excluded patients with a recorded diagnosis for an organic disorder (F00-F09) or who were not residing in Camden or Islington.

We followed patients for up to one year (or until time of death where data was available within EHRs) from their first episode of care with a substance use service. Substance use services included any drug service offered through C&I, which could involve one-to-one key working, day programmes, testing and treatment, assessments, residential detoxification, and rehabilitation centres.

### Measures

#### Outcomes

Our two primary outcomes were (a) rates of re-engagement with C&I substance use services and (b) rates of contact with C&I inpatient/crisis settings. For each patient, we recorded the total count of these outcomes over the one-year follow-up period to estimate rates. Our secondary outcome was time to first contact with C&I crisis or inpatient settings. These included any acute mental health care settings where urgent medical treatment is sought (such as mental health emergency services) and services specifically designed to provide acute mental health crisis support, such as crisis houses and crisis outreach teams.

#### Exposures

Social deprivation was measured using the Index of Multiple Deprivation (IMD) for England (2015) (20), a composite measure of multiple deprivation for small areas based on 37 indicators across 7 domains (income deprivation; employment deprivation; education, skills and training deprivation; health and deprivation and disability; crime; barriers to housing and services; living environment deprivation). Patients were geocoded to the lower super output area (LSOA) of their last recorded place of residence and linked to their IMD quartile (least deprived to most deprived) relative to all LSOA within the catchment area. LSOA information was missing for 17.2% of our study sample, whom we chose to retain as a separate category since we reasoned that a substantial proportion of participants with an opioid-related disorder may have been of no fixed abode at the point of care.

We defined comorbid mental health diagnosis at baseline as a recorded ICD-10 psychiatric diagnosis prior to, or within 30 days of their first episode of care with substance use services. This approach minimised misclassifying psychiatric diagnoses arising subsequent to the substance use disorder, but provided time for follow-up psychiatric assessments to be made immediately after first contact. We initially classified participants as having any of the following comorbid mental and substance use conditions: other substance use (F10, F12-18), common mental disorders (F32-39, F40-49), severe mental illnesses (F20-29, F30, F31, F32.3, F33.3), personality disorders (F60-69), and other mental health conditions (F50, 52, F70-F79, F84.5, F89, F90, F91, F99). From this, we classified participants into four mutually exclusive categories: 1) no comorbid diagnoses; 2) diagnosis in one of these broad sets of psychiatric disorders (but no other non-opioid substance use disorder); 3) diagnosis of a non-opioid substance use disorder (but no other psychiatric disorder); and 4) diagnoses in two or more of these broad sets of comorbid substance use or psychiatric conditions.

#### Confounders

We extracted EHR data on sex, age, ethnicity, and marital status as potential confounders. We also included two additional area-level measures (population density (people per hectare) and social fragmentation index) based on patients’ LSOA. Data for social fragmentation and population density (people per hectare) were estimated using 2011 census data (21), and grouped into quartiles. Consistent with previous studies (22, 23), we estimated social fragmentation as the proportion of unmarried persons, people living at a different address 1 year ago, people living alone, and people privately renting; these data were combined into a single index by summing z-scores for each indicator.

### Analysis

We used Fisher’s exact tests to examine differences in the distribution of categorical exposure and confounder variables between those with and without subsequent mental health service use engagement, and Wilcoxon rank-sum tests for corresponding continuous variables. For our primary outcomes, we used Poisson regression models to compare rates of re-engagement/contact with substance use services and crisis/inpatient settings, respectively. For our secondary outcome, we used Cox proportional hazards regression to model time to first contact with crisis or inpatient settings, and undertook a test of the proportional hazards assumption. Cohort entry was the date of first episode of care with a substance use service, and date of exit was first engagement with a crisis/inpatient service, death (where data was available within HER), or censorship after 365 days. We presented incidence rate ratios (IRR) and Hazard Ratios (HR) for our primary and secondary analyses, respectively, alongside 95% confidence intervals (95% CI). For all analyses, we presented univariable (unadjusted), bivariable (mutually adjusted for the other exposure) and multivariable (fully adjusted for all confounders) results. Data were analysed using Stata 16 (24). Some small cell data (n<10) has been supressed to maintain anonymity.

The study was approved by the Research Database Oversight Committee with ethical approval for epidemiological research granted by the East of England – Cambridge Central Research Ethics Committee (19/EE/0210).

## RESULTS

### Sample characteristics

We identified a total of 768 patients who had a first episode of care with substance use services in Camden and Islington NHS Foundation Trust between 2015 and 2020. Of this sample, 157 (20.4%) patients re-engaged with a substance use service over the following 12 months (Table 1), and 76 (9.90%) patients had contact with crisis or inpatient settings (Table 2).

**Table 1.**
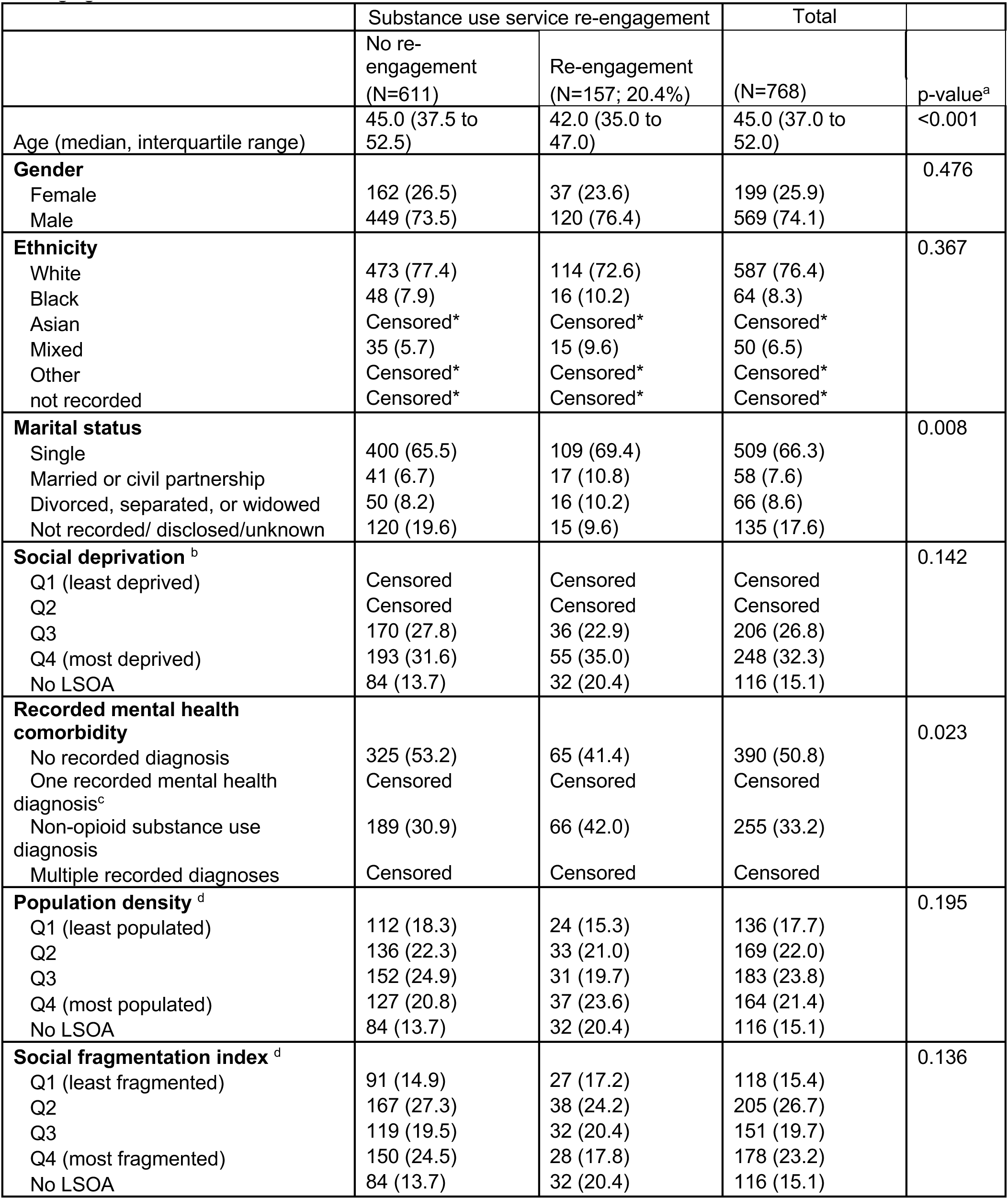
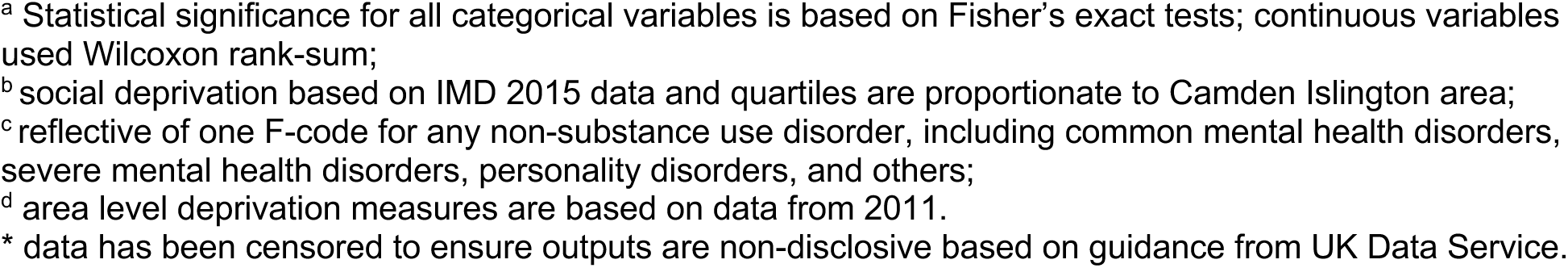
Socio-demographic and clinical characteristics of the total sample in relation to re-engagement with substance use services

|  | Substance use service re-engagement |  | Total |  |
| --- | --- | --- | --- | --- |
|  | No re-engagement<br>(N=611) | Re-engagement<br>(N=157; 20.4%) | (N=768) | p-value<sup>a</sup> |
| Age (median, interquartile range) | 45.0 (37.5 to 52.5) | 42.0 (35.0 to 47.0) | 45.0 (37.0 to 52.0) | <0.001 |
| **Gender** |  |  |  | 0.476 |
| Female | 162 (26.5) | 37 (23.6) | 199 (25.9) |  |
| Male | 449 (73.5) | 120 (76.4) | 569 (74.1) |  |
| **Ethnicity** |  |  |  | 0.367 |
| White | 473 (77.4) | 114 (72.6) | 587 (76.4) |  |
| Black | 48 (7.9) | 16 (10.2) | 64 (8.3) |  |
| Asian | Censored* | Censored* | Censored* |  |
| Mixed | 35 (5.7) | 15 (9.6) | 50 (6.5) |  |
| Other | Censored* | Censored* | Censored* |  |
| not recorded | Censored* | Censored* | Censored* |  |
| **Marital status** |  |  |  | 0.008 |
| Single | 400 (65.5) | 109 (69.4) | 509 (66.3) |  |
| Married or civil partnership | 41 (6.7) | 17 (10.8) | 58 (7.6) |  |
| Divorced, separated, or widowed | 50 (8.2) | 16 (10.2) | 66 (8.6) |  |
| Not recorded/ disclosed/unknown | 120 (19.6) | 15 (9.6) | 135 (17.6) |  |
| **Social deprivation<sup>b</sup>** |  |  |  | 0.142 |
| Q1 (least deprived) | Censored | Censored | Censored |  |
| Q2 | Censored | Censored | Censored |  |
| Q3 | 170 (27.8) | 36 (22.9) | 206 (26.8) |  |
| Q4 (most deprived) | 193 (31.6) | 55 (35.0) | 248 (32.3) |  |
| No LSOA | 84 (13.7) | 32 (20.4) | 116 (15.1) |  |
| **Recorded mental health comorbidity** |  |  |  | 0.023 |
| No recorded diagnosis | 325 (53.2) | 65 (41.4) | 390 (50.8) |  |
| One recorded mental health diagnosis<sup>c</sup> | Censored | Censored | Censored |  |
| Non-opioid substance use diagnosis | 189 (30.9) | 66 (42.0) | 255 (33.2) |  |
| Multiple recorded diagnoses | Censored | Censored | Censored |  |
| **Population density<sup>d</sup>** |  |  |  | 0.195 |
| Q1 (least populated) | 112 (18.3) | 24 (15.3) | 136 (17.7) |  |
| Q2 | 136 (22.3) | 33 (21.0) | 169 (22.0) |  |
| Q3 | 152 (24.9) | 31 (19.7) | 183 (23.8) |  |
| Q4 (most populated) | 127 (20.8) | 37 (23.6) | 164 (21.4) |  |
| No LSOA | 84 (13.7) | 32 (20.4) | 116 (15.1) |  |
| **Social fragmentation index<sup>d</sup>** |  |  |  | 0.136 |
| Q1 (least fragmented) | 91 (14.9) | 27 (17.2) | 118 (15.4) |  |
| Q2 | 167 (27.3) | 38 (24.2) | 205 (26.7) |  |
| Q3 | 119 (19.5) | 32 (20.4) | 151 (19.7) |  |
| Q4 (most fragmented) | 150 (24.5) | 28 (17.8) | 178 (23.2) |  |
| No LSOA | 84 (13.7) | 32 (20.4) | 116 (15.1) |  |

<sup>a</sup> Statistical significance for all categorical variables is based on Fisher's exact tests; continuous variables used Wilcoxon rank-sum;
<sup>b</sup> social deprivation based on IMD 2015 data and quartiles are proportionate to Camden Islington area;
<sup>c</sup> reflective of one F-code for any non-substance use disorder, including common mental health disorders, severe mental health disorders, personality disorders, and others;
<sup>d</sup> area level deprivation measures are based on data from 2011.
\* data has been censored to ensure outputs are non-disclosive based on guidance from UK Data Service.

**Table 2.**
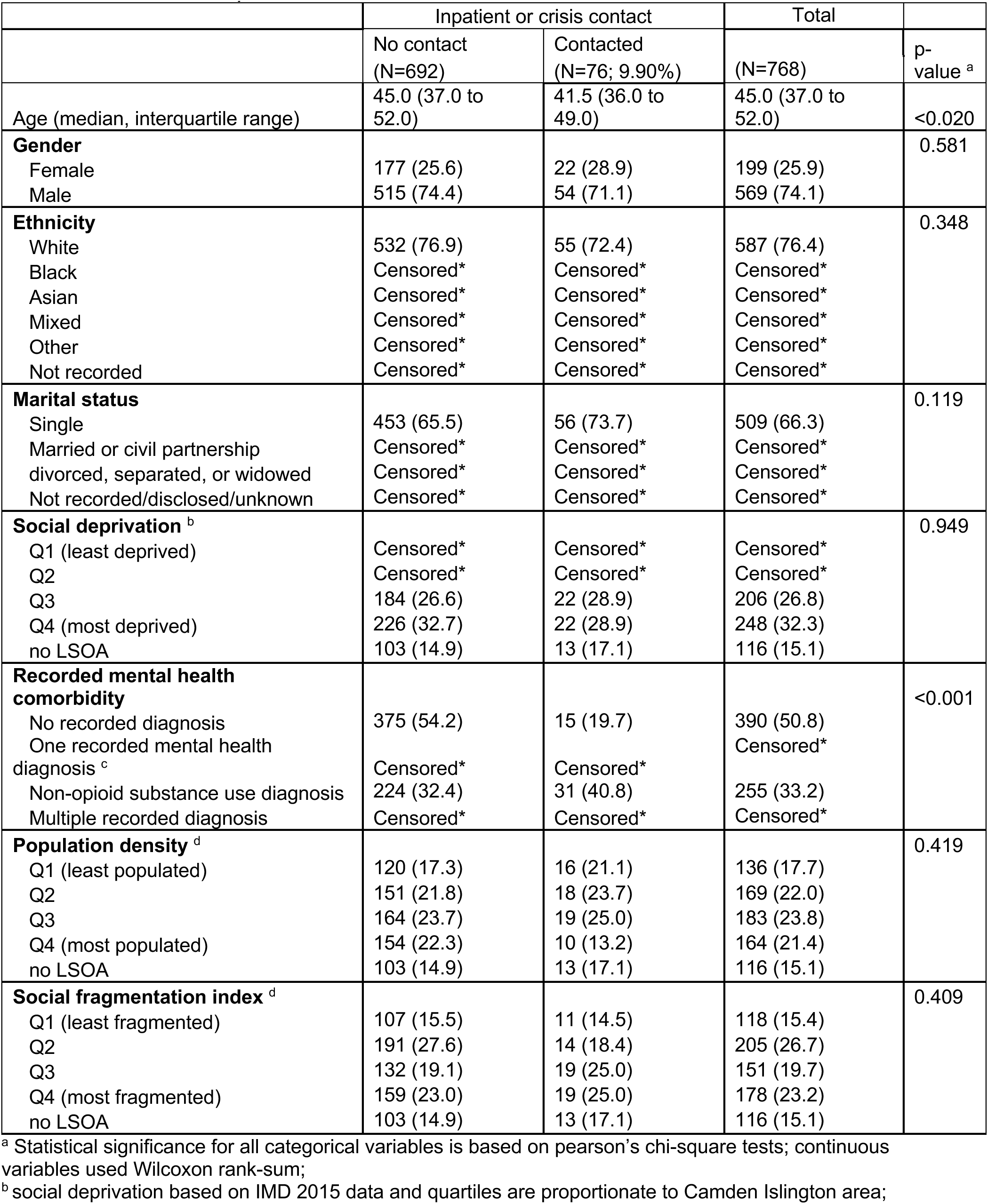
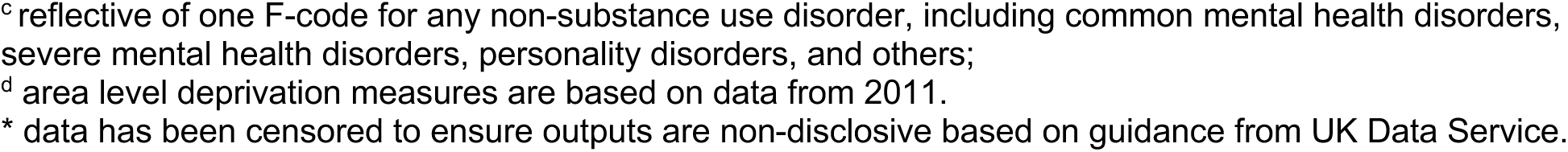
Socio-demographic and clinical characteristics of the total sample in relation to contact with crisis or inpatient services

|  | Inpatient or crisis contact |  | Total |  |
| --- | --- | --- | --- | --- |
|  | No contact<br>(N=692) | Contacted<br>(N=76; 9.90%) | (N=768) | p-value <sup>a</sup> |
| Age (median, interquartile range) | 45.0 (37.0 to 52.0) | 41.5 (36.0 to 49.0) | 45.0 (37.0 to 52.0) | <0.020 |
| **Gender** |  |  |  | 0.581 |
| Female | 177 (25.6) | 22 (28.9) | 199 (25.9) |  |
| Male | 515 (74.4) | 54 (71.1) | 569 (74.1) |  |
| **Ethnicity** |  |  |  | 0.348 |
| White | 532 (76.9) | 55 (72.4) | 587 (76.4) |  |
| Black | Censored* | Censored* | Censored* |  |
| Asian | Censored* | Censored* | Censored* |  |
| Mixed | Censored* | Censored* | Censored* |  |
| Other | Censored* | Censored* | Censored* |  |
| Not recorded | Censored* | Censored* | Censored* |  |
| **Marital status** |  |  |  | 0.119 |
| Single | 453 (65.5) | 56 (73.7) | 509 (66.3) |  |
| Married or civil partnership | Censored* | Censored* | Censored* |  |
| divorced, separated, or widowed | Censored* | Censored* | Censored* |  |
| Not recorded/disclosed/unknown | Censored* | Censored* | Censored* |  |
| **Social deprivation <sup>b</sup>** |  |  |  | 0.949 |
| Q1 (least deprived) | Censored* | Censored* | Censored* |  |
| Q2 | Censored* | Censored* | Censored* |  |
| Q3 | 184 (26.6) | 22 (28.9) | 206 (26.8) |  |
| Q4 (most deprived) | 226 (32.7) | 22 (28.9) | 248 (32.3) |  |
| no LSOA | 103 (14.9) | 13 (17.1) | 116 (15.1) |  |
| **Recorded mental health comorbidity** |  |  |  | <0.001 |
| No recorded diagnosis | 375 (54.2) | 15 (19.7) | 390 (50.8) |  |
| One recorded mental health diagnosis <sup>c</sup> | Censored* | Censored* | Censored* |  |
| Non-opioid substance use diagnosis | 224 (32.4) | 31 (40.8) | 255 (33.2) |  |
| Multiple recorded diagnosis | Censored* | Censored* | Censored* |  |
| **Population density <sup>d</sup>** |  |  |  | 0.419 |
| Q1 (least populated) | 120 (17.3) | 16 (21.1) | 136 (17.7) |  |
| Q2 | 151 (21.8) | 18 (23.7) | 169 (22.0) |  |
| Q3 | 164 (23.7) | 19 (25.0) | 183 (23.8) |  |
| Q4 (most populated) | 154 (22.3) | 10 (13.2) | 164 (21.4) |  |
| no LSOA | 103 (14.9) | 13 (17.1) | 116 (15.1) |  |
| **Social fragmentation index <sup>d</sup>** |  |  |  | 0.409 |
| Q1 (least fragmented) | 107 (15.5) | 11 (14.5) | 118 (15.4) |  |
| Q2 | 191 (27.6) | 14 (18.4) | 205 (26.7) |  |
| Q3 | 132 (19.1) | 19 (25.0) | 151 (19.7) |  |
| Q4 (most fragmented) | 159 (23.0) | 19 (25.0) | 178 (23.2) |  |
| no LSOA | 103 (14.9) | 13 (17.1) | 116 (15.1) |  |

<sup>a</sup> Statistical significance for all categorical variables is based on pearson's chi-square tests; continuous variables used Wilcoxon rank-sum;
<sup>b</sup> social deprivation based on IMD 2015 data and quartiles are proportionate to Camden Islington area;
<sup>c</sup> reflective of one F-code for any non-substance use disorder, including common mental health disorders, severe mental health disorders, personality disorders, and others;
<sup>d</sup> area level deprivation measures are based on data from 2011.
\* data has been censored to ensure outputs are non-disclosive based on guidance from UK Data Service.

Those who re-engaged with substance use services had a lower median age (42.0; IQR: 37.5-52.5) than those who did not (45.0; IQR: 35.0-47.0). Those who re-engaged with substance use services were less likely to have no comorbid diagnosis at first contact (53.2% vs 41.4%; p=0.023). There were differences in marital status although likely associated with the higher rates of no recorded/disclosed marital status for those who did not re-engage with services (19.6% vs 9.6%).

Those who had contact with crisis or inpatient settings during follow-up were also younger than those who did not (median: 41.5 versus 45.0 years; p=0.020; Table 2). We observed differences in comorbidity between those who had and did not have contact with crisis or inpatient settings, such that those who had contact with crisis/inpatient services were substantially less likely to have any comorbid diagnosis than those not in contact with crisis/inpatient services (19.7% versus 54.2%; p<0.001).

### Rates of re-engagement with substance use services

We found no evidence that social deprivation was associated with rates of re-engagement with substance use services during follow-up in our fully-adjusted models, despite some initial evidence that those with no recorded LSOA had higher rates of re-engagement with substance use services in unadjusted (Table 3; IRR: 2.78; 95% CI: 1.28-7.28; p=0.019) and bivariable models (Table 3; IRR: 2.54; 95%CI: 1.16-6.68; p=0.034).

**Table 3.** Re-engagement rates with substance use services in the one-year follow-up period

|  | Unadjusted regression |  |  | Partially adjusted regression <sup>a</sup> |  |  | Fully adjusted regression <sup>b</sup> |  |  |
| --- | --- | --- | --- | --- | --- | --- | --- | --- | --- |
|  | IRR | 95% CI | p-value | IRR | 95% CI | p-value | IRR | 95% CI | p-value |
| **Social deprivation** |  |  |  |  |  |  |  |  |  |
| Q1 (least deprived) | 1 |  |  | 1 |  |  | 1 |  |  |
| Q2 | 1.77 | [0.80,4.67] | 0.198 | 1.65 | [0.75,4.38] | 0.257 | 1.60 | [0.70,4.33] | 0.304 |
| Q3 | 1.49 | [0.68,3.91] | 0.363 | 1.42 | [0.65,3.74] | 0.422 | 1.46 | [0.66,3.90] | 0.392 |
| Q4 (most deprived) | 1.89 | [0.89,4.88] | 0.138 | 1.82 | [0.85,4.71] | 0.163 | 1.93 | [0.65,1.64] | 0.143 |
| no LSOA | 2.78 | [1.28,7.28] | 0.019 | 2.54 | [1.16,6.68] | 0.034 | <sup>c</sup> | <sup>c</sup> | <sup>c</sup> |
| **Recorded mental health comorbidity** |  |  |  |  |  |  |  |  |  |
| No recorded diagnosis | 1 |  |  | 1 |  |  | 1 |  |  |
| One recorded mental health diagnosis | 0.81 | [0.29,1.81] | 0.652 | 0.86 | [0.30,1.92] | 0.742 | 0.67 | [0.16,1.89] | 0.507 |
| Non-opioid substance use diagnosis | 1.43 | [1.05,1.96] | 0.025 | 1.36 | [0.99,1.87] | 0.059 | 1.43 | [0.98,2.08] | 0.064 |
| Multiple recorded diagnosis | 1.26 | [0.78,1.95] | 0.324 | 1.26 | [0.78,1.96] | 0.324 | 1.36 | [0.79,2.23] | 0.244 |

<sup>a</sup> Partially adjusted for the other exposure variable
<sup>b</sup> Fully adjusted for the other exposure and confounders (age, sex, ethnicity, marital status, population density, and social fragmentation)
<sup>c</sup> fully colinear with no LSOA group

There was no evidence that any comorbid mental health or other substance use diagnosis were associated with rates of re-engagement with substance use services during follow-up. Supplement A provides the full set of regression coefficients from these models.

### Rates of contact with crisis/inpatient services

We found no evidence that social deprivation was associated with contact rates with crisis/inpatient services in fully adjusted models (Table 4), despite some initial evidence that those in more deprived quartiles had lower rates contact with crisis/inpatient services that those in the least deprived quartile (Table 4).

**Table 4.** Contact with crisis and inpatient settings in the one-year follow-up period

|  | Unadjusted regression |  |  | Partially adjusted regression <sup>a</sup> |  |  | Fully adjusted regression <sup>b</sup> |  |  |
| --- | --- | --- | --- | --- | --- | --- | --- | --- | --- |
|  | IRR | 95% CI | p-value | IRR | 95% CI | p-value | IRR | 95% CI | p-value |
| **Social deprivation** |  |  |  |  |  |  |  |  |  |
| Q1 (least deprived) | 1 |  |  | 1 |  |  | 1 |  |  |
| Q2 | 0.52 | [0.31,0.90] | 0.017 | 0.45 | [0.26,0.78] | 0.004 | 0.60 | [0.33,1.13] | 0.106 |
| Q3 | 0.72 | [0.45,1.21] | 0.194 | 0.58 | [0.36,0.98] | 0.032 | 0.71 | [0.42,1.25] | 0.219 |
| Q4 (most deprived) | 0.41 | [0.25,0.69] | 0.001 | 0.41 | [0.25,0.71] | 0.001 | 0.66 | [0.36,1.21] | 0.169 |
| no LSOA | 0.76 | [0.45,1.30] | 0.298 | 0.64 | [0.38,1.12] | 0.105 | <sup>c</sup> | <sup>c</sup> | <sup>c</sup> |
| **Recorded mental health comorbidity** |  |  |  |  |  |  |  |  |  |
| No recorded diagnosis | 1 |  |  | 1 |  |  | 1 |  |  |
| One recorded mental health diagnosis | 10.50 | [5.85,18.62] | <0.001 | 9.66 | [5.37,17.18] | <0.001 | 7.02 | [3.63,13.31] | <0.001 |
| Non-opioid substance use diagnosis | 4.53 | [2.95,7.20] | <0.001 | 4.55 | [2.95,7.24] | <0.001 | 3.37 | [2.11,5.53] | <0.001 |
| Multiple recorded diagnosis | 14.03 | [9.20,22.17] | <0.001 | 13.95 | [9.12,22.09] | <0.001 | 11.68 | [7.53,18.71] | <0.001 |

<sup>a</sup> Partially adjusted for the other exposure variable
<sup>b</sup> Fully adjusted for the other exposure and confounders (age, sex, ethnicity, marital status, population density, and social fragmentation)
<sup>c</sup> fully colinear with no LSOA group

In fully adjusted models, any comorbid mental health or other substance use diagnosis was associated with increased rates of contact with crisis/inpatient settings. Those with multiple comorbid mental health diagnoses (IRR: 11.68; 95%CI: 7.53-18.71l; p<0.001) had the highest rates of contact with crisis/inpatient services relative to those without comorbidity. Rates of contact with crisis/inpatient services were also higher among those with only one comorbid mental health disorder (IRR: 7.02; 95%CI: 3.63-13.31; p<0.001) and those with a non-opioid substance use disorder (IRR: 3.37; 95%CI: 2.11-5.53; p<0.001).

Supplement B provides the full set of regression coefficients from these models.

### Time to first contact with crisis/inpatient services

We found no evidence that social deprivation was associated with time to first contact with crisis/inpatient services (Table 5). In contrast, and consistent with rates of crisis/inpatient use above, participants with a comorbid non-opioid substance use disorder (HR: 3.17; 95%CI: 1.68-5.97; p<0.001), or with a single (HR: 7.33; 95%CI: 2.92-18.25; p<0.001) or multiple (HR: 6.95; 95%CI: 3.56-13.57; p<0.001) comorbid mental health diagnoses at baseline had shorter time to first contact with crisis/inpatient services all models. Supplement C provides the full set of regression coefficients from these models.

**Table 5.** Cox regression/survival analysis appearance at crisis/inpatient following access to substance use service

|  | Unadjusted regression |  |  | Partially adjusted regression <sup>a</sup> |  |  | Fully adjusted regression <sup>b</sup> |  |  |
| --- | --- | --- | --- | --- | --- | --- | --- | --- | --- |
|  | HR | 95% CI | p-value | HR | 95% CI | p-value | HR | 95% CI | p-value |
| **Social deprivation** |  |  |  |  |  |  |  |  |  |
| Q1 (least deprived) | 1 |  |  | 1 |  |  | 1 |  |  |
| Q2 | 1.15 | [0.38,3.46] | 0.804 | 1.15 | [0.38,3.46] | 0.804 | 1.21 | [0.37,3.90] | 0.751 |
| Q3 | 1.24 | [0.43,3.59] | 0.694 | 1.24 | [0.43,3.59] | 0.694 | 1.21 | [0.40,3.65] | 0.524 |
| Q4 (most deprived) | 1.01 | [0.35,2.93] | 0.985 | 1.01 | [0.35,2.93] | 0.985 | 1.45 | [0.46,4.54] | 0.524 |
| no LSOA | 1.32 | [0.43,4.06] | 0.624 | 1.32 | [0.43,4.06] | 0.624 | 0.83 | [0.23,2.99] | 0.781 |
| **Recorded mental health comorbidity** |  |  |  |  |  |  |  |  |  |
| No recorded diagnosis | 1 |  |  | 1 |  |  | 1 |  |  |
| One recorded mental health diagnosis | 7.99 | [3.99,18.86] | <0.001 | 7.99 | [3.99,18.86] | <0.001 | 7.33 | [2.92,18.35] | <0.001 |
| Non-opioid substance use diagnosis | 3.31 | [1.79,6.13] | <0.001 | 3.31 | [1.79,6.13] | <0.001 | 3.17 | [1.68,5.97] | <0.001 |
| Multiple recorded diagnosis | 6.84 | [3.55,13.20] | <0.001 | 6.84 | [3.55,13.20] | <0.001 | 6.95 | [3.56,13.57] | <0.001 |

<sup>a</sup> Partially adjusted for the other exposure variable
<sup>b</sup> Fully adjusted for the other exposure and confounders (age, sex, ethnicity, marital status, population density, and social fragmentation)

## DISCUSSION

To our knowledge, this is the first study to use routinely-collected EHRs to investigate social deprivation and comorbid mental health diagnosis as predictors of re-engagement/contact with statutory substance use services, crisis or inpatient settings amongst patients with a diagnosis of opioid use disorder diagnosis following their first episode of treatment. Our study found social deprivation did not influence rates of re-engagement to substance use services up to one year after diagnosis, or rates of contact with crisis/inpatient service use in this period. Our study extends previous knowledge by finding large and robust increases in the rate of contact with crisis/inpatient services, as well as a shorter time to first contact with crisis services, in those with one or more comorbid psychiatric disorders and comorbid non-opioid use substance use disorders at the time of their first episode of care with substance use services.

### Meaning of the findings

Given our findings did not show an influence of deprivation on frequency of re-engagement with substance use services, our results somewhat contrast the majority of studies that find support for increased opioid prescribing for pain treatment in more deprived areas (16, 25–27). One possible explanation is that our catchment area of inner-London is more deprived – on average – than the remainder of England. Further, over 60% of participants with a substance use disorder in our study lived in the most deprived half of Camden and Islington or were missing a residential address, possibly due to homelessness, which is associated with non-medical use of opioids (28, 29). Thus, the absence of differences in rates of service re-engagement and contact by deprivation level in this sample may be due to a floor effect, since most of our sample were already living in very deprived circumstances.

Nearly 50% of our cohort had at least one recorded comorbid mental health or substance use diagnosis alongside their opioid use disorder diagnosis at first contact. This proportion is in-keeping with the published evidence (7–11). Our findings strongly demonstrated that these comorbidities increased rates of contact with crisis/inpatient settings in secondary mental health care for people with an opioid use disorder. Results from a large globally representative, cross-sectional household survey of individuals using opioids found that the presence of at least one comorbid mental health condition (other than an additional substance use disorder) doubled the odds of accessing substance use disorder treatment (30); this aligns with our findings around higher rates and shorter time to contact with crisis/inpatient care in the first 12 months after initial opioid treatment. That study (30), however, did not differentiate between having one, or multiple comorbid mental health diagnoses. Here, we extend the literature to show that people with multiple comorbid mental health diagnoses had the highest rates of contact with crisis and inpatient settings compared with those without any comorbidity. Rates were also high for those with one comorbid mental health or substance use diagnosis, and those with single or multiple mental health comorbidities had the shortest time to crisis/inpatient contact during follow-up.

### Limitations

We used routine data in a secondary mental health care system for a defined catchment area to provide insights into longitudinal predictors of treatment usage for people with an opioid use disorder diagnosis. Using real-world data allowed us to understand the impact of comorbid mental and substance use disorders, as well as deprivation, on longitudinal use of clinical mental health services. Our study also has several limitations. EHR data were primarily collected for clinical purposes. Due to changes in the electronic health care provider contracted by C&I, substance use diagnoses entered in structured fields were not available prior to August 2015, which restricted the case ascertainment period in our study design, even though CRIS records began in 2009. As the end of our study period overlapped with the start of the global COVID-19 pandemic, decreased engagement and contact with health care services could be partially attributable to reductions in service provision and wider alterations to normal health care provision (31). One further limitation of CRIS data is that residential address information was based on the latest clinical record, with historical addresses overwritten by more recent ones. This means that we may have misclassified exposure to deprivation (and other area-level confounders) amongst any patients who moved between LSOA, or became homeless, during their care with C&I (up to 2020). We were unable to assess the extent of this bias in our study, but we cannot exclude reverse causality as an explanation of our results with respect to deprivation. People with opioid use disorders may also be a highly mobile and transient population, which may have led to misclassification of their social deprivation exposure. Further, we did not have information on cohort exit (such as death not recorded within EHRs or emigration from the study region) so were unable to take this into account. Missing data was an issue on some information (notably LSOA and marital status), something which is inherent to most electronic health records. Although we have discussed missingness in LSOA data in terms of homelessness, we cannot exclude the possibility that data were missing for other reasons.

The generalisability of our findings more broadly should be considered. This study only reported on individuals with an opioid use disorder diagnosis who were accessing substance use services through statutory secondary mental health care services. Our results may not generalise to those who solely accessed substance use treatment in their community or through private or third sector organisations, or who had yet to seek any treatment. Additionally, we did not have information on the nature of opioid misuse within our cohort (e.g. prescribed, illicit use). The catchment area of Camden and Islington is also a very densely populated, inner-city area of London, with high levels of deprivation. Our findings may not generalise to other regions of England or elsewhere.

### Implications for policy, practice, and research

Our findings suggest a need to identify comorbid mental health and substance use issues at the first point of treatment and provide appropriate signposting and support to try to reduce the amount of reliance on readmission to the secondary mental health care system. Although due to the size of the sample, we were unable to separate by separate psychiatric conditions, previous research has shown certain psychiatric conditions (such as non-opioid substance use disorders, psychotic diagnosis, and mood disorders) have been found to be more strongly associated with continued opioid use and decreased treatment retention (10, 32). This has implications on care planning and treatment design, for patients with complex needs who are at greater risk of poor treatment outcomes. Interventions and support offered through substance use services could benefit from introducing broader psychiatric and substance use screening, and targeting interventions to ensure individuals are better supported with their additional needs (33), particularly among those presenting with multiple mental health and substance use conditions.

## CONCLUSION

We found that any comorbid mental health condition, including another substance use disorder, was associated with increased rates of contact with crisis/inpatient services among patients within one year of contact with substance use services. By contrast, no association was found between such comorbidities and re-engagement with substance use services. Time to first crisis/inpatient care was at least three times shorter for people with an opioid use disorder diagnosis with a comorbid mental health condition, compared with those without another mental health condition or substance use disorder. We did not observe an association between area-level deprivation and re-engagement/contact with services for individuals with an opioid use disorder diagnosis. Given the common co-occurrence of mental health and substance use disorders among those with an opioid use disorder diagnosis, further observational and interventional work is needed to better understand the needs of this group; allowing them to be better supported in their treatment journeys. Future research should work with people who have lived experience of opioid use to investigate and explore different treatment outcomes, potentially including those accessing services outside of statutory care (such as in third sector organisations).

## Data Availability

Data are owned and controlled by Camden & Islington NHS Foundation Trust. These data are not publicly available in the interest of patient confidentiality and may only be accessed by approved researchers from within a secure firewall (i.e. the data cannot be sent elsewhere), in the same manner as the authors. For more information please contact.

## STATEMENTS AND DECLARATIONS

### Consent to participate

Informed consent was not obtained as data were anonymised.

## REFERENCES

1. United Nations Office on Drugs and Crime. World Drug Report 2022: Booklet 3 Drug Market Trends Cannabis Opioids. United Nations publications; 2022. Available from: https://www.unodc.org/res/wdr2022/MS/WDR22_Booklet_3.pdf

2. The Lancet Public Health. Opioid overdose crisis: time for a radical rethink. Lancet Public Health. 2022;7(3):e195. doi:10.1016/S2468-2667(22)00043-3.

3. Rosner B, Neicun J, Yang JC, Roman-Urrestarazu A. Opioid prescription patterns in Germany and the global opioid epidemic: Systematic review of available evidence. PLoS ONE. 2019;14(8):e0221153. doi:10.1371/journal.pone.0221153.

4. Office for Health Improvement & Disparities. Adult substance misuse treatment statistics 2020 to 2021: report. 2021. Available from: https://www.gov.uk/government/statistics/substance-misuse-treatment-for-adults-statistics-2020-to-2021/adult-substance-misuse-treatment-statistics-2020-to-2021-report

5. Lewis G, Dykxhoorn J, Karlsson H, Khandaker GM, Lewis G, Dalman C, et al. Assessment of the Role of IQ in Associations Between Population Density and Deprivation and Nonaffective Psychosis. JAMA Psychiatry. 2020;77(7):729–36. doi:10.1001/jamapsychiatry.2020.0103

6. Richardson L, Hameed Y, Perez J, Jones PB, Kirkbride JB. Association of Environment With the Risk of Developing Psychotic Disorders in Rural Populations: Findings from the Social Epidemiology of Psychoses in East Anglia Study. JAMA Psychiatry. 2018;75(1):75–83. doi:10.1001/jamapsychiatry.2017.3582

7. Rowe TA, Jacapraro JS, Rastegar DA. Entry into primary care-based buprenorphine treatment is associated with identification and treatment of other chronic medical problems. Addict Sci Clin Pract. 2012;7(1):22. doi:10.1186/1940-0640-7-228.

8. Weinstein ZM, Kim HW, Cheng DM, Quinn E, Hui D, Labelle CT, et al. Long-term retention in Office Based Opioid Treatment with buprenorphine. J Subst Abuse Treat. 2017;74:65–70. doi:10.1016/j.jsat.2016.12.010

9. Simon CB, Tsui JI, Merrill JO, Adwell A, Tamru E, Klein JW. Linking patients with buprenorphine treatment in primary care: Predictors of engagement. Drug Alcohol Depend. 2017;181:58–62. doi: 0.1016/j.drugalcdep.2017.09.017

10. Rosic T, Naji L, Bawor M, Dennis BB, Plater C, Marsh DC, et al. The impact of comorbid psychiatric disorders on methadone maintenance treatment in opioid use disorder: a prospective cohort study. Neuropsychiatr Dis Treat. 2017;13:1399–408. doi:10.2147/NDT.S129480

11. Fareed A, Eilender P, Ketchen B, Buchanan-Cummings AM, Scheinberg K, Crampton K, et al. Factors affecting noncompliance with buprenorphine maintenance treatment. J Addict Med. 2014;8(5):345–50. doi:10.1097/ADM.0000000000000057

12. Burkinshaw P, Knight J, Anders P, Eastwood B, Musto V, White M, et al. An evidence review of the outcomes that can be expected of drug misuse treatment in England. London: Public Health England; 2017.

13. Mattick RP, Breen C, Kimber J, Davoli M. Methadone maintenance therapy versus no opioid replacement therapy for opioid dependence. Cochrane Database of Systematic Reviews. 2003;2.Art. No.: CD002209. doi:10.1002/14651858.CD002209.

14. Hooker SA, Sherman MD, Lonergan-Cullum M, Sattler A, Liese BS, Justesen K, et al. Mental Health and Psychosocial Needs of Patients Being Treated for Opioid Use Disorder in a Primary Care Residency Clinic. J Prim Care Community Health. 2020;11:2150132720932017.

15. Macfarlane GJ, Beasley M, Jones GT, Stannard C. The epidemiology of regular opioid use and its association with mortality: Prospective cohort study of 466 486 UK biobank participants. EClinicalMedicine. 2020;21. doi:10.1016/j.eclinm.2020.100321

16. Nowakowska M, Zghebi SS, Perisi R, Chen L-C, Ashcroft DM, Kontopantelis E. Association of socioeconomic deprivation with opioid prescribing in primary care in England: a spatial analysis. Journal of Epidemiology and Community Health. 2020:jech-2020-214676. doi:10.1136/jech-2020-214676

17. Cantone RE, Garvey B, O’Neill A, Fleishman J, Cohen D, Muench J, et al. Predictors of Medication-Assisted Treatment Initiation for Opioid Use Disorder in an Interdisciplinary Primary Care Model. J Am Board Fam Med. 2019;32(5):724–31. doi:10.3122/jabfm.2019.05.190012

18. Morton CM. Community social deprivation and availability of substance use treatment and mutual aid recovery groups. Subst Abuse Treat Prev Policy. 2019;14(1):33. doi:10.1186/s13011-019-0221-6

19. Werbeloff N, Osborn DPJ, Patel R, Taylor M, Stewart R, Broadbent M, et al. The Camden & Islington Research Database: Using electronic mental health records for research. PLOS ONE. 2018;13(1):e0190703. doi: 10.1371/journal.pone.0190703

20. Ministry of Housing, Communities & Local Government. English indices of deprivation 2015. 2015. Available from: https://www.gov.uk/government/statistics/english-indices-of-deprivation-2015

21. Official census and labour market statistics. 2011 Census. 2011. Available from: https://www.nomisweb.co.uk/sources/census_2011.

22. Allardyce J, Gilmour H, Atkinson J, Rapson T, Bishop J, McCreadie R. Social fragmentation, deprivation and urbanicity: relation to first-admission rates for psychoses. The Br J Psychiatry. 2005;187(5):401–6. doi:10.1192/bjp.187.5.401

23. Kirkbride JB, Jones PB, Ullrich S, Coid JW. Social Deprivation, Inequality, and the Neighborhood-Level Incidence of Psychotic Syndromes in East London. Schizophr Bull. 2012;40(1):169–80. doi:10.1093/schbul/sbs151

24. StataCorp. Stata statistical software: release 16. College Station, TX: StataCorp LLC; 2019.

25. Chen TC, Chen LC, Kerry M, Knaggs RD. Prescription opioids: Regional variation and socioeconomic status - evidence from primary care in England. Int J Drug Policy. 2019;64:87–94. doi:10.1016/j.drugpo.2018.10.013

26. Curtis HJ, Croker R, Walker AJ, Richards GC, Quinlan J, Goldacre B. Opioid prescribing trends and geographical variation in England, 1998-2018: a retrospective database study. Lancet Psychiatry. 2019;6(2):140–50. doi:10.1016/S2215-0366(18)30471-1

27. Mordecai L, Reynolds C, Donaldson LJ, de CWAC. Patterns of regional variation of opioid prescribing in primary care in England: a retrospective observational study. Br J Gen Pract. 2018;68(668):e225–e33. doi:10.3399/bjgp18X695057

28. Yamamoto A, Needleman J, Gelberg L, Kominski G, Shoptaw S, Tsugawa Y. Association between homelessness and opioid overdose and opioid-related hospital admissions/emergency department visits. Soc Sci Med. 2019;242:112585. doi:10.1016/j.socscimed.2019.112585

29. Milaney K, Passi J, Zaretsky L, Liu T, O’Gorman CM, Hill L, et al. Drug use, homelessness and health: responding to the opioid overdose crisis with housing and harm reduction services. Harm Reduct J. 2021;18(1):92. doi:10.1186/s12954-021-00539-8

30. Harris MG, Bharat C, Glantz MD, Sampson NA, Al-Hamzawi A, Alonso J, et al. Cross-national patterns of substance use disorder treatment and associations with mental disorder comorbidity in the WHO World Mental Health Surveys. Addiction. 2019;114(8):1446–59. doi:10.1111/add.14599.

31. Alexander K, Pogorzelska-Maziarz M, Gerolamo A, Hassen N, Kelly EL, Rising KL. The impact of COVID-19 on healthcare delivery for people who use opioids: a scoping review. Subst Abuse Treat Prev Policy. 2021;16(1):60. doi:10.1186/s13011-021-00395-6.

32. Bharat C, Larney S, Barbieri S, Dobbins T, Jones NR, Hickman M, et al. The effect of person, treatment and prescriber characteristics on retention in opioid agonist treatment: a 15-year retrospective cohort study. Addiction. 2021;116(11):3139–52. doi:10.1111/add.15514

33. Griffin ML, Dodd DR, Potter JS, Rice LS, Dickinson W, Sparenborg S, et al. Baseline characteristics and treatment outcomes in prescription opioid dependent patients with and without co-occurring psychiatric disorder. Am J Drug Alcohol Abuse. 2014;40(2):157–62. doi:10.3109/00952990.2013.842241.

