## Supplemental tables for "Investigating social deprivation and comorbid mental health diagnosis as predictors of treatment access among patients with an opioid use disorder using substance use services: a prospective cohort study"

### SUPPLEMENTAL MATERIAL

*Supplement Table A: Full Poisson regressions for re-engagement rates with substance use services in the one-year follow-up period*

| | Unadjusted regression | | Partially adjusted regression [a] | | Fully adjusted regression [b] | |
|---|---|---|---|---|---|---|
| | IRR | 95% CI | IRR | 95% CI | IRR | 95% CI |
| Age | 0.98** | [0.96,0.99] | | | 0.97** | [0.96,0.99] |
| **Gender** | | | | | | |
| Female | 1 | | | | 1 | |
| Male | 1.11 | [0.80,1.58] | | | 1.14 | [0.77,1.71] |
| **Ethnicity** | | | | | | |
| White | 1 | | | | 1 | |
| Black | 1.21 | [0.72,1.93] | | | 1.12 | [0.58,1.98] |
| Asian | 0.83 | [0.29,1.82] | | | 0.86 | [0.26,2.12] |
| Mixed | 1.47 | [0.85,2.36] | | | 1.09 | [0.55,1.97] |
| Other | 0.72 | [0.25,1.58] | | | 0.82 | [0.24,2.09] |
| **Marital status** | | | | | | |
| single | 1 | | | | 1 | |
| married | 1.22 | [0.72,1.93] | | | 1.44 | [0.77,2.50] |
| divorced, separated, or widowed | 1.13 | [0.67,1.78] | | | 1.48 | [0.83,2.48] |
| not disclosed/recorded or unknown | 0.49** | [0.29,0.79] | | | 0.52* | [0.27,0.92] |
| **Social deprivation** | | | | | | |
| Q1 (least deprived) | 1 | | 1 | | 1 | |
| Q2 | 1.77 | [0.80,4.67] | 1.65 | [0.75,4.38] | 1.60 | [0.70,4.33] |
| Q3 | 1.49 | [0.68,3.91] | 1.42 | [0.65,3.74] | 1.46 | [0.66,3.90] |
| Q4 (most deprived) | 1.89 | [0.89,4.88] | 1.82 | [0.85,4.71] | 1.93 | [0.65,1.64] |
| no LSOA | 2.78* | [1.28,7.28] | 2.54* | [1.16,6.68] | [c] | [c] |
| **Recorded mental health comorbidity** | | | | | | |
| No recorded diagnosis | 1 | | 1 | | 1 | |
| One recorded mental health diagnosis | 0.81 | [0.29,1.81] | 0.86 | [0.30,1.92] | 0.67 | [0.16,1.89] |
| Non-opioid substance use diagnosis | 1.43* | [1.05,1.96] | 1.36 | [0.99,1.87] | 1.43 | [0.98,2.08] |
| Multiple recorded diagnosis | 1.26 | [0.78,1.95] | 1.65 | [0.75,4.38] | 1.36 | [0.79,2.23] |
| **Social fragmentation index** | | | | | | |
| Q1 (least deprived) | 1 | | | | 1 | |
| Q2 | 0.83 | [0.52,1.33] | | | 0.68 | [0.42,1.11] |
| Q3 | 0.94 | [0.58,1.53] | | | 0.81 | [0.81,1.35] |
| Q4 (most deprived) | 0.73 | [0.44,1.20] | | | 0.65 | [0.38,1.10] |
| no LSOA | 1.42 | [0.89,2.29] | | | [c] | [c] |
| **Population Density** | | | | | | |
| Q1 (least deprived) | 1 | | | | 1 | |
| Q2 | 1.19 | [0.85,1.66] | | | 1.15 | [0.81,1.63] |
| Q3 | 1.39* | [1.01,1.92] | | | 1.45* | [1.03,2.05] |
| Q4 (most deprived) | 1.31 | [0.95,1.81] | | | 1.33 | [0.93,1.90] |
| no LSOA | 1.65** | [1.19,2.28] | | | [c] | [c] |

Exponentiated coefficients; 95% confidence intervals in brackets
[*] $p < 0.05$, [**] $p < 0.01$, [***] $p < 0.001$

PREPRINT

[a] Partially adjusted regressions were adjusted for using exposure variables (social deprivation and recorded mental health comorbidity); [b] fully adjusted regressions adjusted for exposure and confounders (age, sex, ethnicity, marital status, population density, and social fragmentation); [c] colinear with no LSOA group.

PREPRINT

*Supplement Table B: Full Poisson regressions for contact with crisis and inpatient settings in the one year follow-up period*

| | Unadjusted regression | | Partially adjusted regression [a] | | Fully adjusted regression [b] | |
|---|---|---|---|---|---|---|
| | IRR | 95% CI | IRR | 95% CI | IRR | 95% CI |
| Age | 0.97$^{***}$ | [0.96,0.99] | | | 0.97$^{**}$ | [0.96,0.99] |
| **Gender** | | | | | | |
| Female | 1 | | | | 1 | |
| Male | 0.74$^{*}$ | [0.55,0.99] | | | 0.89 | [0.64,1.26] |
| **Ethnicity** | | | | | | |
| White | 1 | | | | 1 | |
| Black | 0.66 | [0.35,1.13] | | | 0.43$^{*}$ | [0.18,0.87] |
| Asian | 0.94 | [0.40,1.85] | | | 0.72 | [0.25,1.67] |
| Mixed | 0.98 | [0.54,1.62] | | | 0.93 | [0.50,1.60] |
| Other | 1.16 | [0.58,2.09] | | | 1.34 | [0.55,2.91] |
| **Marital status** | | | | | | |
| single | 1 | | | | 1 | |
| married | 0.31$^{**}$ | [0.12,0.64] | | | 0.43 | [0.16,0.94] |
| divorced, separated, or widowed | 0.91 | [0.55,1.41] | | | 1.03 | [0.56,1.76] |
| not disclosed/recorded or unknown | 0.33$^{***}$ | [0.19,0.54] | | | 0.50$^{*}$ | [0.28,0.84] |
| **Social deprivation** | | | | | | |
| Q1 (least deprived) | 1 | | 1 | | 1 | |
| Q2 | 0.52$^{*}$ | [0.31,0.90] | 0.45$^{**}$ | [0.26,0.78] | 0.60 | [0.33,1.13] |
| Q3 | 0.72 | [0.45,1.21] | 0.58$^{*}$ | [0.36,0.98] | 0.71 | [0.42,1.25] |
| Q4 (most deprived) | 0.41$^{***}$ | [0.25,0.69] | 0.41$^{***}$ | [0.25,0.71] | 0.66 | [0.36,1.21] |
| no LSOA | 0.76 | [0.45,1.30] | 0.64 | [0.38,1.12] | [c] | [c] |
| **Recorded mental health comorbidity** | | | | | | |
| No recorded diagnosis | 1 | | 1 | | 1 | |
| One recorded mental health diagnosis | 10.50$^{***}$ | [5.85,18.62] | 9.66$^{***}$ | [5.37,17.18] | 7.02$^{***}$ | [3.63,13.31] |
| Non-opioid substance use diagnosis | 4.53$^{***}$ | [2.95,7.20] | 4.55$^{***}$ | [2.95,7.24] | 3.37$^{***}$ | [2.11,5.53] |
| Multiple recorded diagnosis | 14.03$^{***}$ | [9.20,22.17] | 13.95$^{***}$ | [9.12,22.09] | 11.68$^{***}$ | [7.53,18.71] |
| **Social fragmentation index** | | | | | | |
| Q1 (least fragmented) | 1 | | | | 1 | |
| Q2 | 0.65 | [0.42,1.01] | | | 0.53$^{**}$ | [0.33,0.85] |
| Q3 | 0.74 | [0.47,1.17] | | | 0.65 | [0.40,1.07] |
| Q4 | 0.94 | [0.62,1.44] | | | 0.66 | [0.42,1.07] |
| no LSOA | 1.07 | [0.69,1.67] | | | [c] | [c] |
| **Population Density** | | | | | | |
| Q1 (least pop) | 1 | | | | 1 | |
| Q2 | 0.74 | [0.51,1.07] | | | 0.73 | [0.48,1.09] |
| Q3 | 0.48$^{***}$ | [0.32,0.72] | | | 0.68 | [0.42,1.09] |
| Q4 | 0.28$^{***}$ | [0.17,0.46] | | | 0.38$^{***}$ | [0.21,0.65] |
| no LSOA | 0.79 | [0.53,1.18] | | | [c] | [c] |

Exponentiated coefficients; 95% confidence intervals in brackets

$^{*}$ $p < 0.05$, $^{**}$ $p < 0.01$, $^{***}$ $p < 0.001$

[a] Partially adjusted regressions were adjusted for using exposure variables (social deprivation and recorded mental health comorbidity); [b] fully adjusted regressions adjusted for exposure and confounders

PREPRINT

(age, sex, ethnicity, marital status, population density, and social fragmentation); [c] colinear with no LSOA group.

PREPRINT

*Supplement C: Cox regressions for contact with crisis and inpatient settings in the one year follow-up and tests of proportional hazards assumption*

| | Unadjusted regression | | Partially adjusted regression [a] | | Fully adjusted regression [b] | |
|---|---|---|---|---|---|---|
| | HR | 95% CI | HRR | 95% CI | HR | 95% CI |
| Age | 0.97* | [0.95,1.00] | | | 0.97 | [0.95,1.00] |
| **Gender** | | | | | | |
| Female | 1 | | | | 1 | |
| Male | 0.86 | [0.52,1.40] | | | 0.78 | [0.46,1.30] |
| **Ethnicity** | | | | | | |
| White | 1 | | | | 1 | |
| Black | 0.83 | [0.33,2.06] | | | 0.73 | [0.29,1.86] |
| Asian | 1.72 | [0.62,4.76] | | | 1.38 | [0.46,4.09] |
| Mixed | 1.58 | [0.72,3.47] | | | 1.23 | [0.55,2.78] |
| Other | 1.91 | [0.76,4.77] | | | 2.45 | [0.87,6.94] |
| **Marital status** | | | | | | |
| single | 1 | | | | 1 | |
| married | 0.62 | [0.22,1.71] | | | 0.53 | [0.18,1.56] |
| divorced, separated, or widowed | 1.28 | [0.63,2.58] | | | 1.75 | [0.83,3.67] |
| not disclosed/recorded or unknown | 0.46 | [0.21,1.01] | | | 0.47 | [0.21,1.07] |
| **Social deprivation** | | | | | | |
| Q1 (least deprived) | 1 | | 1 | | 1 | |
| Q2 | 1.15 | [0.38,3.46] | 1.15 | [0.38,3.46] | 1.21 | [0.37,3.90] |
| Q3 | 1.24 | [0.43,3.59] | 1.24 | [0.43,3.59] | 1.21 | [0.40,3.65] |
| Q4 (most deprived) | 1.01 | [0.35,2.93] | 1.01 | [0.35,2.93] | 1.45 | [0.46,4.54] |
| no LSOA | 1.32 | [0.43,4.06] | 1.32 | [0.43,4.06] | 0.83 | [0.23,2.99] |
| **Recorded mental health comorbidity** | | | | | | |
| No recorded diagnosis | 1 | | 1 | | 1 | |
| One recorded mental health diagnosis | 7.99*** | [3.99,18.86] | 7.99*** | [3.99,18.86] | 7.33*** | [2.92,18.35] |
| Non-opioid substance use diagnosis | 3.31*** | [1.79,6.13] | 3.31*** | [1.79,6.13] | 3.17*** | [1.68,5.97] |
| Multiple recorded diagnosis | 6.84*** | [3.55,13.20] | 6.84*** | [3.55,13.20] | 6.95*** | [3.56,13.57] |
| **Social fragmentation index** | | | | | | |
| Q1 (least fragmented) | 1 | | | | 1 | |
| Q2 | 0.71 | [0.32,1.56] | | | 0.46 | [0.20,1.05] |
| Q3 | 1.38 | [0.66,2.90] | | | 1.07 | [0.49,2.34] |
| Q4 | 1.15 | [0.55,2.41] | | | 0.81 | [0.36,1.79] |
| no LSOA | 1.22 | [0.55,2.73] | | | c | c |
| **Population Density** | | | | | | |
| Q1 (least pop) | 1 | | | | 1 | |
| Q2 | 0.90 | [0.46,1.76] | | | 0.75 | [0.37,1.54] |
| Q3 | 0.87 | [0.45,1.70] | | | 0.88 | [0.42,1.87] |
| Q4 | 0.50 | [0.23,1.11] | | | 0.51 | [0.22,1.17] |
| no LSOA | 0.96 | [0.46,2.00] | | | c | c |

Exponentiated coefficients; 95% confidence intervals in brackets

* $p < 0.05$, ** $p < 0.01$, *** $p < 0.001$

[a] Partially adjusted regressions were adjusted for using exposure variables (social deprivation and recorded mental health comorbidity); [b] fully adjusted regressions adjusted for exposure and confounders

PREPRINT

(age, sex, ethnicity, marital status, population density, and social fragmentation); [c] colinear with no LSOA group.

PREPRINT
